# Early joint attention abilities measured by the ADOS-2 predict subsequent expressive language development in minimally verbal autistic children

**DOI:** 10.1101/2025.09.07.25335088

**Authors:** Tanya Nitzan, Moran Bachrach, Michal Ilan, Michal Faroy, Danel Waissengreen, Analya Michaelovsky, Dikla Zagdon, Yair Sadaka, Omer Bar Yosef, Ditza Zachor, Einat Avni, Idan Menashe, Gal Meiri, Judah Koller, Ilan Dinstein

## Abstract

**Background:** Most preschool autistic children exhibit substantial language delays, yet only ∼25% remain minimally verbal (MV) throughout life. Previous studies have demonstrated that development of expressive language abilities is crucial for improving long-term outcomes. This study aimed to identify early predictors of later expressive language development specifically in MV preschool autistic children.

**Methods:** We analyzed prospective data collected from 99 MV autistic children, who were 27.7 months old at diagnosis, on average. All children completed an ADOS-2 assessment at diagnosis and again 12-24 months later. We classified children into three expressive language groups at follow up: MV, one-word phase, and phrases phase. Logistic regression analyses were used to identify significant predictors of expressive language abilities at follow up. Predictors included ADOS-2 social affect (SA) calibrated severity scores (CSS), ADOS-2 restricted and repetitive behavior (RRB) CSS, cognitive scores, and joint attention (JA) scores, derived from a summation of six ADOS-2 items.

**Results:** Children who successfully developed expressive language abilities at follow-up (i.e., were in the one-word or phrases groups) had significantly lower ADOS-2 SA CSS and JA scores at diagnosis. Logistic regression analyses demonstrated that both ADOS-2 SA CSS and JA scores at diagnosis predicted expressive language outcomes while cognitive scores and ADOS-2 RRB CSS did not. Moreover, concurrent improvements in JA were significantly larger in children who developed expressive language.

**Conclusions:** Preschool MV autistic children with better social abilities, and specifically JA abilities, at diagnosis were more likely to develop expressive language within 1-2 years. JA scores derived from the ADOS-2 offer an easily accessible and widely available measure with important prognostic value for these children.

## Introduction

Language delays are the most common initial concern that motivates parents of autistic children to seek clinical advice (Nitzan et al., 2023) and parents commonly rate language acquisition as a top priority for early intervention (Coonrod & Stone, 2004). Approximately 50–60% of autistic children exhibit severe language delays and are minimally verbal (MV) at the age of three (Kissine et al., 2023). The definition of MV varies between studies, ranging from children who do not speak at all to those who use less than 50 words (Koegel et al., 2020). Despite intense intervention, only half of MV children will develop expressive language abilities by the age of seven, with the other half remaining MV throughout life, corresponding to ∼25% of the entire autism population (Fountain et al., 2012; Pickles et al., 2014; Rose et al., 2016; Tager-flusberg & Kasari, 2013; Weismer & Kover, 2015). Of those who develop expressive language abilities between the ages of three to seven, various language trajectories are observed, ranging from gradual to unexpectedly rapid language gains at different ages (Anderson et al., 2007; Baghdadli et al., 2012; Fountain et al., 2012; Georgiades et al., 2022; Pickles et al., 2014; Weismer & Kover, 2015; Wodka et al., 2013). Most importantly, the emergence of expressive language is associated with the development of higher cognitive abilities, better adaptive functioning, and reduced severity of core autism symptoms, leading to better long-term life outcomes (Mouga et al., 2020) including wellbeing and mental health (Brignell et al., 2018; Howlin et al., 2013). There is, therefore, strong motivation to identify early predictors of expressive language development specifically for MV autistic children and develop new interventions for those who are unlikely to acquire expressive language using existing intervention protocols.

Previous studies have reported mixed findings regarding early child characteristics that may predict later expressive language development in MV autistic children. Examined predictors have included the overall severity of autism symptoms (Weismer & Kover, 2015), social abilities (Latrèche et al., 2024; Thurm et al., 2015; Weismer & Kover, 2015; Yoder et al., 2015; Zachor et al., 2007), cognitive abilities (Anderson et al., 2007; Kilili-Lesta et al., 2024; Mouga et al., 2020; Norrelgen et al., 2015; Thurm et al., 2015; Weismer & Kover, 2015), and joint attention (JA) abilities (Anderson et al., 2007; Kasari et al., 2023; Kilili-Lesta et al., 2024; McDaniel et al., 2017; Sandbank et al., 2017; Yoder et al., 2015). Note that we are focusing this introduction on longitudinal studies that specifically examined language development in MV autistic children (i.e., those with severe language delays), rather than studies that included autistic children with mild language delays.

Several studies have examined ADOS-2 or older ADOS scores as potential predictors of expressive language development in MV preschool autistic children and reported mixed findings. One study reported a significant relationship between early total ADOS Calibrated Severity Scores (CSS) and later expressive language scores (Weismer & Kover, 2015).

However, others did not, whether using total ADOS-2 CSS (Thurm et al., 2015), ADOS-2 social affect (SA) CSS (Latrèche et al., 2024; Thurm et al., 2015), or ADOS social communication scores (Yoder et al., 2015). ADOS-2 restricted and repetitive behaviors (RRB) CSS also did not predict later expressive language development in MV preschool autistic children (Thurm et al., 2015).

One important aspect of early social communication is JA, where two individuals allocate their attention to the same object, action, or target in a synchronized manner, enabling them to communicate effectively (Mundy, 2018; Mundy et al., 2016). The ability of a child to respond to JA bids from adults seems to be particularly important for learning the meaning of specific words (Bottema-Beutel, 2016) and by extension, for developing language abilities. Different studies have quantified JA abilities in different ways. While some used manual coding of videotaped interactions during the Early Social Communication Scales (ESCS) (Kasari et al., 2023; McDaniel et al., 2017; Saul & Norbury, 2020) or the Communication and Symbolic Behavior Scales Developmental Profile Behavior Sample (CSBS) (McDaniel et al., 2017; Sandbank et al., 2017; Saul & Norbury, 2020; Yoder et al., 2015), others used scores from specific items of the ADOS (Anderson et al., 2007) or parent questionnaires (Kilili-Lesta et al., 2024). Most studies reported that different measures of early JA behaviors including child initiated JA (IJA) and responding to JA (RJA) are associated with better expressive language outcomes at later ages (Anderson et al., 2007; Kasari et al., 2023; Kilili-Lesta et al., 2024; McDaniel et al., 2017; Sandbank et al., 2017; Yoder et al., 2015), while a few studies did not (McDaniel et al., 2017; Saul & Norbury, 2020).

Similarly, most studies have reported that higher cognitive abilities at early ages are associated with greater likelihood of subsequent language acquisition (Anderson et al., 2007; Kilili-Lesta et al., 2024; Mouga et al., 2020; Norrelgen et al., 2015; Thurm et al., 2015), potentially outperforming autism severity measures as a predictor of language development (Thurm et al., 2015; Weismer & Kover, 2015). However, other studies did not find early cognitive ability to predict later expressive language development (Latrèche et al., 2024; Yoder et al., 2015).

The goal of the current study was to revisit this topic with a relatively large longitudinal sample of Israeli preschool autistic children who were defined as MV at diagnosis using strict criteria (i.e., children who do not speak at all). We prospectively followed these children over a one-to-two-year period as they participated in repeated standardized testing with ADOS-2 assessments, allowing us to compare the initial characteristics of those who successfully acquired basic language abilities with those who remained MV.

## Methods

### Participants and procedures

Our sample of convenience included 99 children (82 boys), 18-50 months old at diagnosis, who were diagnosed with autism between 2016 and 2022, and did not have any known genetic disorders. All children completed ADOS-2 (Lord et al., 2012) assessments at two time points, one at diagnosis (T1) and another 12–24 months later (2T), and were classified as MV based on their score on the A1 item of the ADOS-2 at diagnosis (Table 1). This sample was extracted from the National Autism Database of Israel (Dinstein et al., 2020), managed by the Azrieli National Centre for Autism and Neurodevelopment Research (ANCAN). ANCAN is a collaboration between Ben-Gurion University of the Negev (BGU) and 8 clinical sites throughout Israel where children with autism are diagnosed and followed up prospectively. This study was approved by the SUMC Helsinki Committee.

**Table 1:** Characteristics of the children (all MV at T1), separated into three subgroups according to their verbal abilities at T2.

|  | **MV**<br>**(n=49)** | **One-word**<br>**(n=33)** | **Phrases**<br>**(n=17)** | **Overall**<br>**(n=99)** |
| --- | --- | --- | --- | --- |
| **Measures** | **Mean (SD)** |  |  |  |
| Age at diagnosis (M) | 28.6 (7.31) | 27.0 (7.22) | 26.6 (5.80) | 27.7 (7.03) |
| Age at follow-up (M) | 42.9 (7.25) | 40.8 (7.67) | 44.2 (6.70) | 42.4 (7.34) |
| Sex, % boys | 81.6 | 84.8 | 82.4 | 82.8 |
| Paternal age at diagnosis (Y) | 38.5 (7.25) | 35.0 (7.64) | 36.1 (6.32) | 36.9 (7.34) |
| Paternal education (Y) | 12.7 (2.28) | 13.1 (2.36) | 12.8 (1.79) | 12.8 (2.22) |
| Maternal age at diagnosis (Y) | 35.6 (5.46) | 31.8 (5.73) | 33.2 (4.60) | 33.9 (5.63) |
| Maternal education (Y) | 13.2 (2.29) | 13.1 (1.96) | 14.2 (2.30) | 13.3 (2.20) |
| Verbal ability on | 6.47 (0.50) | 4.58 (0.50) | 2.71 (0.470) | 5.19 (1.50) |

|  | MV<br>(n=49) | One-word<br>(n=33) | Phrases<br>(n=17) | Overall<br>(n=99) |
| --- | --- | --- | --- | --- |
| Measures | Mean (SD) |  |  |  |
| ADOS-2 at T2 |  |  |  |  |
| ADOS-2 SA-CSS at T1 | 8.94 (1.48) | 8.00 (1.89) | 7.47 (2.24) | 8.37 (1.84) |
| ADOS-2 RRB-CSS at T1 | 7.53 (1.57) | 7.24 (1.35) | 6.94 (1.52) | 7.33 (1.49) |
| ADOS-2 CSS at T1 | 8.67 (1.30) | 7.85 (1.73) | 7.12 (1.93) | 8.13 (1.66) |
| Cognitive score at T1 | 72.2 (13.3) | 73.4 (10.7) | 75.5 (15.9) | 73.2 (12.9) |
| JA score at T1 | 10.8 (1.42) | 9.70 (2.08) | 8.71 (2.73) | 10.1 (2.06) |
| ADOS-2 SA-CSS at T2 | 7.39 (1.77) | 6.27 (2.14) | 5.35 (2.60) | 6.67 (2.18) |
| ADOS-2 RRB-CSS at T2 | 8.22 (1.21) | 8.09 (1.49) | 8.12 (1.54) | 8.16 (1.35) |
| ADOS-2 CSS at T2 | 7.84 (1.68) | 7.03 (2.04) | 6.06 (2.44) | 7.26 (2.03) |
| JA score at T2 | 9.96 (2.02) | 6.64 (2.99) | 3.53 (2.55) | 7.75 (3.45) |
| Duration in special education (M) | 7.43 (5.60) | 7.91 (5.73) | 7.65 (7.47) | 7.63 (5.94) |

ADOS-2: Autism Diagnostic Observation Schedule; CSS: calibrated severity scores; SA: social affect; RRB: restricted and repetitive behaviors, JA: JA score, SE: Special Education, M: Months, Y: Years.

### Measures

#### ADOS-2

Children completed the Toddler module or modules 1–3 of the ADOS-2 (Lord et al., 2012) according to their age and verbal abilities. All ADOS-2 assessments were administered by experienced clinicians with research reliability. ADOS-2 Calibrated Severity Scores (CSS) allow comparison of autism severity across children of different ages and language capabilities and across time points(Esler et al., 2015; Gotham et al., 2009) and were also computed separately for social affect (SA) and restricted and repetitive behaviors (RRBs) (Esler et al., 2015; Hus et al., 2014).

#### Non-verbal cognitive abilities

76 children (76.8% of the sample) completed cognitive testing at T1 and missing scores were imputed (see below). Cognitive ability was measured using different assessments according to chronological and mental age, as determined by a licensed psychologist. The Bayley Scales of Infant and Toddler Development, Third Edition (Vanegas, 2021) was used at T1 with ∼90.8% children, the Mullen Scales of Early Learning (MSEL; Mullen, 1995) was used with ∼6.6% children, and the Wechsler Preschool and Primary Scale of Intelligence, Third Edition (Luiselli et al., 2013) was used at T1 with ∼2.6% children. The three tests yield equivalent standardized scores with a mean of 100 and a standard deviation of 15. Since strong correlations exist between the Bayley and Wechsler tests, as well as between the MSEL and Bayley tests (Lense et al., 2014; Vanegas, 2021), we combined scores across tests in our analysis.

#### Verbal abilities

Verbal ability was evaluated using the A1 item of the ADOS-2 assessment, which evaluates the spoken language abilities of the child. In module 1 and the toddler module, this item scores verbal abilities from no spoken language to using pairs of words, in module 2 from single words to combinations of three or more words, and in module 3 from combinations of 2–3 words to complex sentences. We used the coding system developed by Visser et al. (2017) (Visser et al., 2017) to create a common 8-point spoken language scale across modules with the following values:

0. Children use sentences in a largely correct fashion (complex utterances with >2 clauses).
1. Children exhibit relatively complex speech (occasional utterances with >2 clauses) with recurrent grammatical errors.
2. Children exhibit non-echoed speech with utterances of >3 words.
3. Children mainly use individual 2-to 3-word phrases, with or without minimal grammar.
4. Children mainly use individual words with occasional simple phrases.
5. Children only use individual words (minimum of 5 different words).
6. Children only use echoed speech (<5 words).
7. No language production at all.

We then classified the sample into three verbal ability groups at follow-up (T2): MV (scores 6-7), one-word phase (scores 4-5), and phrases (scores 2-3). No children in our sample displayed fluent speech (i.e., scores 0-1) at T2.

#### Joint attention

JA abilities were estimated by summing 6 items from the ADOS-2: Pointing (item A7 in the Toddlers module and Module 1, item A6 in Module 2), Gesturing (item A8 in the Toddlers module and Module 1 and item A7 in Module 2), Showing (item B12 in the Toddlers module, B9 in Module1, and B5 in Module 2), Initiating JA (item B13 in the Toddlers module, B10 in Module1, and B6 in Module 2), Unusual Eye contact (item B1 in all modules), and Reaction to JA (item B14 in the Toddlers module, B11 in Module1, and B7 in Module 2). In line with the ADOS algorithm, scores of 3 were changed to 2, and scores of 8 were changed to 0, yielding a total JA scale of 0-12 with higher scores indicating poorer JA abilities (i.e., larger difficulties).

Our estimation of JA abilities differed slightly from a that of others who based it on a factor analysis of the ADOS algorithm (Gotham et al., 2007, 2008; Oosterling et al., 2010). These studies reported that one of the factors identified in an exploratory factor analysis included the following five items: pointing, gesturing, showing, initiating JA, and unusual eye contact in Module 1 (Some Words) and Module 2. In Module 1 (No Words) the factor differed and included gesturing, showing, initiating JA, response to JA, and unusual eye contact (i.e., in this module the factor included the response to JA item instead of pointing). Since both pointing and response to JA are items that clearly measure JA abilities, and to consistently quantify JA across children who completed different modules, we decided to sum the scores of all six items regardless of module. Note that previous research with typically developing children has reported that pointing facilitates JA development, elicits contingent parental speech (McGillion et al., 2013; Rollins, 2003; Tomasello et al., 2007) and is critical for early language acquisition (Watson et al., 2013). This further motivated the inclusion of this item for all children.

#### Additional variables

Parents completed follow-up questionnaires at T2 where they reported maternal and paternal age at time of their child’s diagnosis, their years of education, and the duration of time that their child had attended special education up to T2. The number of respondents varied by variable, with paternal age reported for 87 children (87.9% of the sample), paternal education for 80 children (80.8 of the sample), maternal age for 94 children (94.9% of the sample), maternal education for 88 children (88.9% of the sample), and the Duration in special education for 93 children (93.9% of the sample). Missing data for the remaining children were imputed (see below).

### Data analysis and statistics

All statistical analyses were conducted using RStudio (RStudio Inc., Boston, MA), version 4.4.3. All children included in the study were minimally verbal (MV) at diagnosis (T1) and classified into three groups according to their verbal ability score at follow-up (T2): MV (scored 6/7), one-word phase (scored 4/5), or able to use phrases (scored 3 or less). We imputed missing data, including cognitive scores and parent follow-up questionnaire responses, using the Random Forests technique with 20 iterations as implemented in the *mice* package in RStudio (van Buuren & Groothuis-Oudshoorn, 2011). We then used the imputed dataset in all analyses. A series of one-way analyses of covariance (ANCOVAs) were conducted to determine whether initial cognitive, ADOS-2 SA-CSS, ADOS-2 RRB-CSS, or JA scores at T1 differed significantly across verbal ability groups as defined at T2. We controlled for multiple covariates including age of diagnosis, maternal education, parental age at diagnosis, duration of time in special education, and duration of time between ADOS-2 evaluations. Post hoc pairwise comparisons were conducted using Tukey’s Honestly Significant Difference (HSD) test to adjust for multiple comparisons following significant effects in the ANCOVA. Eta squared (η²) was calculated for each ANCOVA to assess the effect size of group differences. Equivalent analyses were performed to compare longitudinal changes in ADOS-2 and JA scores across the three verbal ability groups.

We also performed several logistic regression analyses to predict verbal ability at T2 using multiple T1 measures as predictors. Model fit was assessed using McFadden’s pseudo R² and the Akaike Information Criterion (AIC). In all analyses, statistical significance was set at α = 0.05.

## Results

We first performed ANCOVA analyses to assess whether ADOS-2 SA CSS, ADOS-2 RRB CSS, cognitive scores, or JA scores at diagnosis (T1) differed across children who developed language abilities or remained MV at follow up (T2). These analyses (Figure 1) revealed significant differences in ADOS-2 SA CSS at T1 across the three verbal ability groups (F(2, 90) = 5.73, p =.005, η² = 0.10). Post hoc Tukey HSD comparisons showed that children in the MV group had significantly higher SA CSS than those in the phrases group (mean difference =-1.47, 95% CI [-2.59,-0.34], p =.007) or one-word group (mean difference =-0.94, 95% CI [-1.84,-0.04], p =.039). There were no significant differences between the one-word and phrases groups (mean difference =-0.53, 95% CI [-1.72, 0.66], p =.543) and there were no significant relationships with any of the covariates (maternal education, parental age, duration in special education, and duration between ADOS-2 assessments), except for age of diagnosis (F(1, 90) = 13.50, p <.001). This demonstrated that MV autistic children with lower ADOS-2 SA CSS at diagnosis were more likely to develop expressive language abilities at T2 even when accounting for their age of diagnosis.

**Figure 1:**
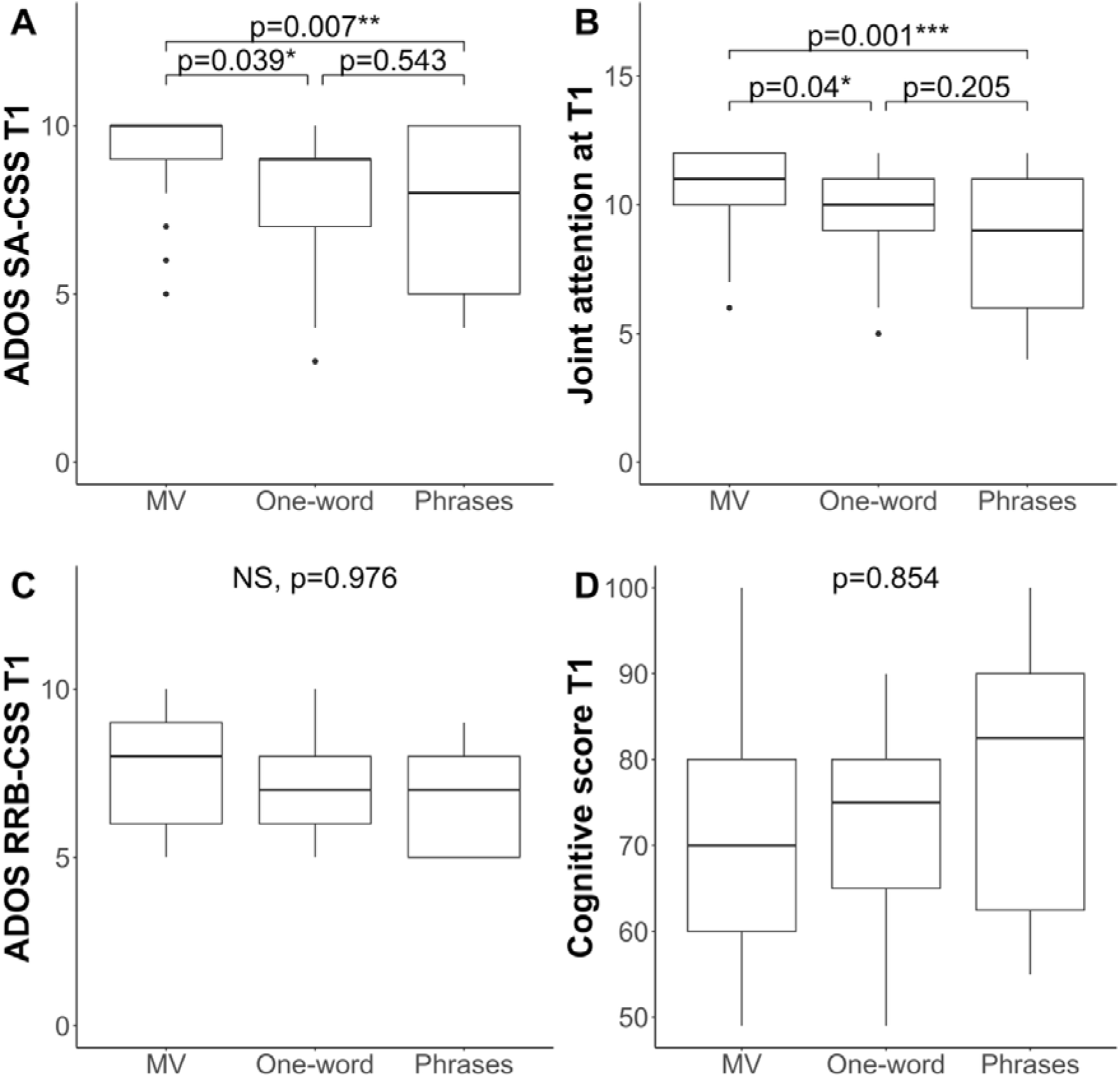
Box and whisker plots demonstrate T1 differences across children who remained MV versus those who developed expressive language abilities (one-word or phrases) at T2. (A) ADOS-2 SA-CSS (B) Joint attention (C) ADOS-2 RRB-CSS (D) Cognitive Scores. Statistical significance of differences across groups from ANCOVA analyses are presented in each panel.

**Figure 2:**
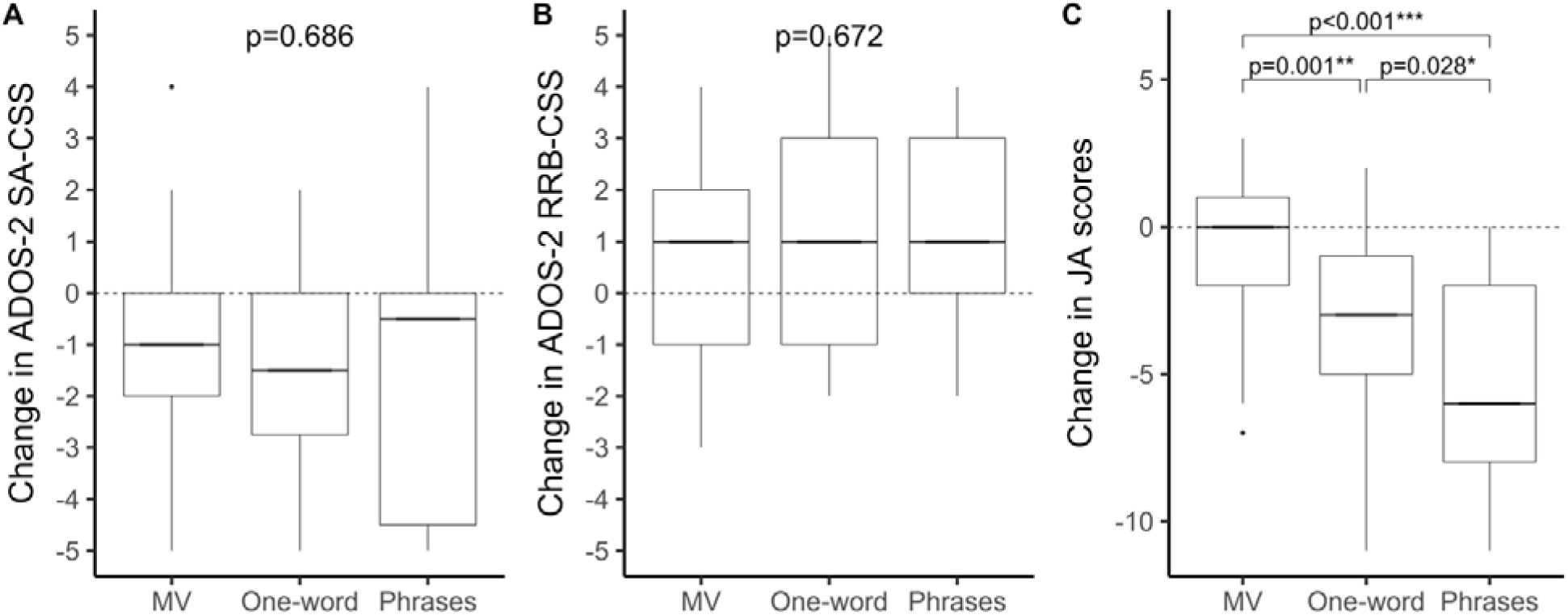
Box and whisker plots of longitudinal changes in ADOS-2 scores (SA, RRB, JA) across verbal abilities groups. (A) Change in ADOS-2 SA-CSS (T1 - T2). Note that negative values indicate improvements (i.e. reduction in ADOS-2 scores) over time. (B) Change in ADOS-2 RRB-CSS. (C) Change in JA score. Horizontal lines represent the group median; no-change is represented by the dashed line.

JA scores at diagnosis (T1) also differed significantly across the three verbal ability groups (F(2, 90) = 6.311, p =.003, η² = 0.15). Post hoc Tukey HSD indicated that children in the MV group had significantly higher JA scores than those in the phrases group (mean difference =-2.07, 95% CI [-3.37,-0.77], p =.0007), and also significantly higher scores than the one-word group (mean difference =-1.08, 95% CI [-2.12,-0.04], p =.040). There were no significant differences between the one-word and phrases groups (mean difference =-0.99, 95% CI [-2.37, 0.39], p =.205). All covariates were not significantly associated with language outcome. Hence, MV autistic children with lower JA scores at T1 were more likely to develop expressive language abilities at T2 even when accounting for their age of diagnosis.

There were no significant differences in cognitive scores (F(2, 90) = 0.15, p =.854) or ADOS-2 RRB CSS (F(2, 90) = 0.02, p =.976) at diagnosis across the three verbal ability groups.

Next, we performed two logistic regression analyses (Table 2) to evaluate the ability of combinations of T1 measures to predict verbal ability at T2. The first logistic regression model included ADOS-2 SA CSS, ADOS-2 RRB CSS, cognitive score, age of diagnosis, maternal age at diagnosis, paternal age at diagnosis, maternal education, duration in special education, and duration of time between T1 and T2 (i.e., diagnosis and follow up). In this analysis ADOS-2 SA CSS at T1 and maternal age at diagnosis emerged as significant predictors of verbal ability at T2. In a second logistic regression analysis we replaced ADOS-2 SA CSS with JA scores and included all other predictors as described above. In this analysis, JA scores and maternal age at diagnosis emerged as significant predictors of verbal abilities at T2. Note that replacing SA CSS with JA scores yielded a model with similar predictive power as indicated by similar pseudo R^2^ (0.187 versus 0.198 respectively) and AIC values (131.61 versus 130.03 respectively). Moreover, SA CSS and JA scores exhibited similar odds ratios (0.41 and 0.44) within their respective models demonstrating similar predictive power.

**Table 2:** Logistic Regression analyses for predicting verbal ability at T2.

| Model 1 |  |  |  |  |  | Model 2 |  |  |  |  |  |
| --- | --- | --- | --- | --- | --- | --- | --- | --- | --- | --- | --- |
| Variable | OR | SE | 95% CI | z | p | Variable | OR | SE | 95% CI | z | p |
| Intercept | 1.00 | 0.00 | 1.00, 1.00 | 2.16 | 0.031* | Intercept | 1.00 | 0.00 | [1.00, 1.00] | 2.23 | 0.027* |
| **ADOS-2 SA CSS** | **0.41** | **0.30** | **[0.22, 0.71]** | **-2.96** | **0.003\*\*** | **JA** | **0.38** | **0.31** | **[0.19, 0.67]** | **-3.09** | **0.002\*\*** |
| ADOS-2 RRB CSS | 1.04 | 0.26 | [0.62, 1.75] | 0.14 | 0.892 | ADOS-2 RRB CSS | 1.08 | 0.27 | [0.64, 1.84] | 0.28 | 0.779 |
| Cognitive score | 0.94 | 0.26 | [0.56, 1.55] | -0.26 | 0.797 | Cognitive score | 0.87 | 0.27 | [0.51, 1.47] | -0.52 | 0.601 |
| Age of diagnosis | 0.62 | 0.28 | [0.35, 1.04] | -1.76 | 0.079 | Age of diagnosis | 0.69 | 0.26 | [0.41, 1.15] | -1.39 | 0.166 |
| Maternal education | 1.50 | 0.27 | [0.89, 2.62] | 1.51 | 0.132 | Maternal education | 1.66 | 0.28 | [0.98, 2.95] | 1.83 | 0.067 |
| **Maternal age at diagnosis** | **0.50** | **0.35** | **[0.24, 0.97]** | **-2.00** | **0.045\*** | **Maternal age at diagnosis** | **0.46** | **0.35** | **[0.22, 0.90]** | **-2.20** | **0.028\*** |
| Paternal age at diagnosis | 0.88 | 0.32 | [0.46, 1.66] | -0.41 | 0.684 | Paternal age at diagnosis | 0.90 | 0.32 | [0.48, 1.71] | -0.32 | 0.748 |
| Duration in special education | 1. | 0. | [0. , 2. ] | 1.37 | 0.172 | Duration in special education | 1. | 0.2 | [0. , . ] | 1.28 | 0.200 |
| Duration between T1 and T2 | 0. | 0.28 | [0.5 , 1. ] | -0.36 | 0.719 | Duration between T1 and T2 | 0. | 0.2 | [0.5 , 1. ] | -0.40 | 0.689 |
| Observations: 99 |  |  |  |  |  | Observations: 99 |  |  |  |  |  |
| Pseudo R-squared (McFadden): 0.18 |  |  |  |  |  | Pseudo R-squared (McFadden): 0.198 |  |  |  |  |  |
| AIC: 131.61 |  |  |  |  |  | AIC: 130.03 |  |  |  |  |  |
| *$p < 0.05$ | | | | | | | | | | | |

### Longitudinal changes in core autism symptoms across language groups

Additional one-way ANCOVA tests demonstrated that longitudinal changes in ADOS-2 SA-CSS between T1 and T2 did not differ significantly across verbal ability groups (F(2, 90) = 0.38, p =.686) nor did longitudinal changes in ADOS-2 RRB-CSS (F(2, 90) = 0.40, p =.672). In contrast, longitudinal changes in JA scores did differ significantly across verbal ability groups (F(2, 90) = 18.14, p <.001, η² = 0.28). Post hoc Tukey HSD comparisons showed that children in the MV group exhibited smaller gains (i.e., reductions in JA scores) than children in the one-word group (M difference =-2.24, 95% CI [-3.70, - 0.79], p =.001) or the phrases group (M difference =-4.36, 95% CI [-6.18,-2.54], p <.001). Significant difference was also found between the one-word and phrases groups (M difference =-2.11, 95% CI [-4.05,-0.18], p = 0.03). All covariates were not significantly associated with language outcome. Hence, MV autistic children who develop expressive language abilities also develop better JA abilities even when accounting for their age of diagnosis.

## DISCUSSION

Our results demonstrate that early ADOS-2 SA CSS and JA scores are similarly predictive of successful expressive language development in young MV autistic children 1-2 years after diagnosis. This was evident in similar effect sizes for SA (OR = 0.41) and JA (OR = 0.38) predictors in their respective logistic regression models (Table 2). Since JA scores were extracted from a subset of ADOS-2 SA items, this suggests that they contain the essential information needed to predict subsequent expressive language development. In addition, JA scores improved significantly with time in MV autistic children who developed expressive language while SA CSS did not. This suggests that there is a particularly strong relationship between JA ADOS-2 items and expressive language development.

### JA scores and later expressive language development in MV children

Multiple previous studies have already reported that better JA skills at diagnosis were associated with greater development of expressive language skills in MV autistic children (Anderson et al., 2007; Kasari et al., 2023; Kilili-Lesta et al., 2024; McDaniel et al., 2017; Sandbank et al., 2017; Yoder et al., 2015). However, many of these studies used manual coding of videotaped interactions during the Early Social Communication Scales (ESCS) (Kasari et al., 2023; McDaniel et al., 2017; Saul & Norbury, 2020) or the Communication and Symbolic Behavior Scales Developmental Profile Behavior Sample (CSBS) (McDaniel et al., 2017; Sandbank et al., 2017; Saul & Norbury, 2020; Yoder et al., 2015), which are time-consuming and unscalable. An important contribution of the current study is in demonstrating that JA scores, extracted from six items of the ADOS-2 can predict expressive language development in MV autistic children.

Previous studies using ADOS/ADOS-2 items to estimate JA abilities (Anderson et al., 2007; Harrison et al., 2016; Maljaars et al., 2012; Nowell et al., 2018; Sano et al., 2021; Thurm et al., 2007) have used different subsets of 2-5 items of the six JA factor items identified by Gotham et al., (2007, 2008). In the current study we included all six JA items regardless of the ADOS-2 module completed by the child to measure JA abilities in an equivalent manner across all children (see Methods).

Of the studies that estimated JA abilities from ADOS item scores, only two used such estimates to predict language development in MV autistic children. Anderson et al., (2007) summed the scores of four ADOS items: gestures, pointing, showing, and spontaneous initiation of JA (children with some speech) or gestures, use of other’s body to communicate, response to name, and response to JA (nonverbal children). Thurm et al., (2007) used scores from the initiation of JA and response to JA items separately. In both cases JA scores predicted the development of expressive language in MV autistic children at follow-up. Our findings further support the conclusion that JA scores extracted from the ADOS-2 are useful for predicting expressive language development in MV autistic children and propose an easy to implement calculation with strong predictive power.

### Early ADOS-2 SA scores and later expressive language development in MV children

Despite the widespread use of the ADOS and ADOS-2 assessments in autism research, relatively few studies have examined their utility in predicting expressive language development of MV children and have yielded mixed results (Latrèche et al., 2024; Thurm et al., 2015; Weismer & Kover, 2015; Yoder et al., 2015). Importantly, two key studies that used the newer ADOS-2 test and extracted SA CSS reported no association with later expressive language development in 47 (Thurm et al., 2015) and 44 (Latrèche et al., 2024) children who were MV at the age of ∼3.5 and 2.5 years-old, respectively. Our results did show a significant association between early ADOS-2 SA CSS and later expressive language abilities. This difference across studies may be due to several reasons.

First, one important difference between our study and both the Thurm et al., (2015) and Latrèche et al., (2024) studies was in how language groups were defined. Our MV criteria were the strictest, such that all children in the current study had no language or only echoed speech at diagnosis. In contrast, Thurm et al., (2015) included children who spoke some single words and occasional phrases in their MV group. Latrèche et al., (2024) used a parent-reported questionnaire to estimate language abilities, performed a clustering analysis, and defined MV as the cluster with expressive language scores that were 4 standard deviations below population standards (i.e., included children who spoke single words and occasional phrases according to parent report). Second, there were differences in the samples recruited for the different studies. Children in the Thurm et al., (2015) study were severely impacted individuals with cognitive scores of ∼60, on average, while the Latrèche et al., (2024) and our study included children with cognitive scores of ∼70, on average, which are more representative of the general autism population (Dinstein et al., 2020). We also did not include any children with known genetic disorders, a criterion that was not reported by the previous studies. Third, While the Thurm et al., (2015) study used ADOS assessments with children diagnosed according to DSM-4 criteria, our study and the Latrèche et al., (2024) study used ADOS-2 assessments with children diagnosed according to DSM-5 criteria.

Finally, the sample size in our study was twice the size of the previous ones, thereby affording considerably larger statistical power.

Hence, in contrast to the previous studies, our study compared strictly defined MV children who did develop expressive language with those who did not, while using a large sample of ADOS-2 assessments collected from community-recruited idiopathic autism cases and diagnosed according to DSM-5 criteria. We, therefore, believe that our findings are persuasive in demonstrating that early ADOS-2 SA CSS are associated with later expressive language outcomes in this specific subgroup of MV autistic children.

### Other predictors of expressive language development in MV children

Previous studies have reported that early cognitive abilities are a significant predictor of later expressive language development in MV autistic children (Anderson et al., 2007; Kilili-Lesta et al., 2024; Latrèche et al., 2024; Mouga et al., 2020; Thurm et al., 2015) while others have not (Latrèche et al., 2024; Yoder et al., 2015). Our findings did not demonstrate such an association despite our relatively large sample, which had larger statistical power than most previous studies. There may be several reasons for these differences across studies. First, some of the studies relied on parent reports to estimate autism severity and/or language abilities (Anderson et al., 2007; Kilili-Lesta et al., 2024; Latrèche et al., 2024; Mouga et al., 2020), which may introduce differences in reporter bias. Second, the type of assessment used to estimate cognitive abilities differed across studies. We used the nonverbal subscale of the Bayley-III at Time 1, excluding all language-dependent items, while others used total cognitive scores that include language-dependent items, which are inappropriate for assessing the cognitive abilities of MV children (Anderson et al., 2007; Ellis Weismer et al., 2010; Yoder et al., 2015). Nevertheless, Thurm et al., (2015), did report that a nonverbal cognitive score (Mullen NVDQ) predicted expressive language development in their sample. Hence, there remains some controversy regarding the potential ability of non-verbal cognitive scores to predict later expressive language development in MV autistic children In contrast, there seems to be a consensus that the severity of early RRB symptoms is not associated with later expressive language development in MV autistic children. This was clear in our results as well as in those of multiple previous longitudinal studies (Bal et al., 2020; Ray-Subramanian & Susan Ellis Weismer, 2012; Thurm et al., 2015; Troyb et al., 2016). Consequently, these findings suggest that RRB behaviors do not interfere with the acquisition of expressive language in MV autistic children.

### Limitations

We note several limitations in the current study. First, we did not have any information regarding the types, duration, and intensity of interventions received by individual children. While we did have parent reports about the duration the child had spent in a special education setting, this measure does not necessarily correspond to a specific intervention approach or estimate its intensity. Hence, there may have been dramatic differences in the amount of intervention received by individual children that were not accounted for in our analyses. Second, all our predictive measures were entirely based on clinical observations using the ADOS-2 and Bayley III. Adding parental reported measures would have been informative in determining the relative predictive strength of parent reported measures versus clinical observation measures. Third, our study lacked a formal standardized language assessment that would have been important for assessing more refined expressive language changes over time.

## Conclusions

Our findings demonstrate that preschool MV autistic children with better social abilities and particularly JA abilities are more likely to succeed in developing expressive language regardless of their initial non-verbal cognitive abilities or RRB symptom severity. We show that JA scores from the ADOS-2 items specified by pointing, gesturing, showing, unusual eye contact, initiating JA, and reacting to JA, offer a simple, widely available JA measure that is predictive of later language development. Finally, our findings highlight the need to develop new interventions for MV preschool autistic children with poor initial social and JA abilities who are less likely to acquire spoken language with existing interventions.

## Data Availability

De-identified data used in all analyses will be made available from the corresponding author upon reasonable request.

